# A method for prioritizing risk groups for early SARS-CoV-2 Vaccination, By the Numbers

**DOI:** 10.1101/2020.12.18.20248504

**Authors:** Clement J. McDonald, Seo H. Baik, Zhaonian Zheng, Liz Amos

## Abstract

**Background:** Given the limited supply of two COVID-19 vaccines, it will be important to choose which risk groups to prioritize for vaccination in order to get the most health benefits from that supply.

**Method:** In order to help decide how to get the maximum health yield from this limited supply, we implemented a logistic regression model to predict COVID-19 death risk by age, race, and sex and did the same to predict COVID-19 case risk.

**Results:** Our predictive model ranked all demographic groups by COVID-19 death risk. It was highly concentrated in some demographic groups, e.g. 85+ year old Black, Non-Hispanic patients suffered 1,953 deaths per 100,000. If we vaccinated the 17 demographic groups at highest COVID-19 death ranked by our logistic model, it would require only 3.7% of the vaccine supply needed to vaccinate all the United States, and yet prevent 47% of COVID-19 deaths. Nursing home residents had a higher COVID-19 death risk at 5,200 deaths/100,000, more than our highest demographic risk group. Risk of prison residents and health care workers (HCW) were lower than that of our demographic groups with the highest risks.

We saw much less concentration of COVID-19 case risk in any demographic groups compared to the high concentration of COVID-19 death in some such groups. We should prioritize vaccinations with the goal of reducing deaths, not cases, while the vaccine supply is low.

**Conclusion:** SARS-CoV-2 vaccines protect against severe COVID-19 infection and thus against COVID-19 death per vaccine studies. Allocating at least some of the early vaccine supplies to high risk demographic groups could maximize lives saved. Our model, and the risk estimate it produced, could help states define their vaccine allocation rules.

## Introduction

As of December 14, SARS-CoV-2 had infected more than 16 million persons and caused more than 298,000 deaths^1^ in the U.S. It has infected and killed ten times the number in the 2018 flu epidemic (35.5 million cases and 34,200 deaths^2^) and nearly 100 times the number in the 1952 polio epidemic (57,879 cases and 3,145 deaths^3^).

Two COVID-19 vaccines with 95% efficacy and no serious adverse events^4^ have Food and Drug Administrative (FDA) Emergency Use Authorization. However, vaccine supply is limited and may remain so for many months. Given the increasing number of cases and death, methods for prioritizing the risk groups who will receive the most benefit from the available supply are important. In this report, we propose a method to numerically rank the vaccination benefit per demographic risk group.

## Methods

### Two data sets for defining COVID-19 risk based on demographic categories

We used provisional monthly death count data from the National Center for Health Statistics (NCHS)^5^ to create a logistic regression model for predicting death rates by demographic category. NCHS’s internal system carries records about deaths due to 14 selected conditions including COVID-19, coded to ICD-10 code U07.1. This information is reported grouped by age range, race/ethnicity, and sex. It includes no individual level data. We only used COVID death data from this source. It was dated November 25, 2020 and included COVID-19 death information from April 1 (when the COVID-19 ICD code was created) through October 30, 2020. Age ranges begin at 0-4 years and increase in 10-year increments until age 84, beyond which all cases are lumped in one, 85+ group. Cells containing fewer than 10 individuals are flagged as such and delivered with no content.

We used data from the Centers for Disease Control and Prevention (CDC)’s COVID-19 Surveillance Public Use Data^6^, updated December 4, 2020 with data up to November 19, 2020 to create a logistic regression model for predicting case rates. It carries patient level data including age group, race, ethnicity, sex, hospitalization, death status and other variables. Ages are binned differently from the NCHS data. Age ranges start at 0-9 and step in 10-year intervals until 70-79; above which all records are classed as 80+ years. We grouped the individual records from this data set into combinations of age range, sex, and racial-ethnic categories.

In both data sets, race-ethnicity is combined into five terms using categories defined by the 2019 NCHS Bridged-Race Population Estimates^7^: American Indian/Alaska Native, Non-Hispanic; Asian, Non-Hispanic; Black, Non-Hispanic; Native Hawaiian/Other Pacific Island, Non-Hispanic, Multiple/Other, Non-Hispanic; and Hispanic/Latino. To simplify the tables and the text, we use “Black” to mean Black, Non-Hispanic”, Hispanic to mean Hispanic/Latino, “American Indian” to mean American Indian/Alaska Native, Non-Hispanic, “Asian” to mean, Asian, plus Native Hawaiian/Other Pacific Islander, Non-Hispanic, and “White” to mean White, Non-Hispanic. We will use “race” to mean the combined race-ethnicity terms.

We obtained estimated population counts from the 2019 NCHS Bridged-Race Population Estimates^7^ for each demographic category in each of the data sets, as approximates for 2020 counts, and used them to compute death and case rates per 100,000 U.S. inhabitants for each demographic category. We only used de-identified and publicly available data in this study.

### Data about special risk groups

We obtained information about nursing home cases and deaths from the Centers for Medicare & Medicaid Service (CMS).^8^ It reports COVID-19 cases and deaths per nursing home, without patient level data or demographic breakdown. The total number of nursing home residents (1.2 million) came from Kaiser Family Foundation’s analysis.^9^

We obtained information about health care workers from several incomplete sources.^10,11^ For example, one source with information on 11.9 million COVID-19 cases lacked health care worker status for 80%.^11^

Prison popoulation was estimated from the Prison Policy Initative^12^ and the prison COVID death rates came from the COVID-19 Behind Bars project.^13^

#### Statistical methods

To provide a simple overview of the death and case rates by demographic categories, we generated cross-tabulations by race, age, and sex for both data sets. These tables include roll ups that provide the number of the deaths and cases by race, sex, and age alone as well as by pairwise combination of these three variables.

We used logistic regression analysis^14^ to create predictive models with COVID death and case rates, as their respective outcomes. Because the race data was absent in a large proportion the CDC surveillance records, we ran the analysis of case rates two ways: one parallel to the death analysis which used the subset of CDC surveillance records that carried complete age, sex, and race data and another that included almost all of the surveillance data but ignored race.

Logistic regression produces an equation based on characteristics (variables) on a training set of cases with known outcomes. The equation will then calculate the probability of that outcome for a new case when fed the same set of variables. Logistic regression has the advantage that it can be used on grouped data or on individual level data, can accommodate continuous as well as categorical variables, and provides confidence limits on its estimates. It was necessary to include all pair wise interactions between age, sex, and race to obtain stable statistic in these analyses.

The input to for these analyses were tables with a row for each demographic category e.g. age >80, sex = male, race = Black. Each row also carries a count of individuals within category who suffered the outcome (a COVID-19 death or a COVID-19 case) and a count of U.S. inhabitants within that category from NCHS Bridged-Race Population.^7^ The output of this analysis was the respective likelihood of a COVID-19 death or COVID-19 case for each demographic category.

SARS-CoV-2 vaccine prevents severe cases and thus death.^4^ We assume vaccination of a given risk group will prevent 100% of the COVID-19 deaths predicted for that group. For simplicity sake, we also presume it will prevent 100% of the cases, a slight exaggeration over the reported 95% effectiveness.

Because our goal is to develop a prioritization algorithm to save the maximum number of lives, we counted all deaths equally. Other weightings, such as life years lost are possible, but the WHO SAGE report^15^ argues for use of a simple unweighted count, as we did. We used SAS Version 9.4 PROC FREQ to generate the cross-tabulation breakdowns and SAS Version 9.4 PROC LOGISTIC to generate the logistic regressions.

## RESULTS

### Break down of death rates

The NCHS data set dated November 25, 2020^5^ carried results for 220,582 COVID-19 deaths occurring between April 1 and October 30. No age or race data was missing but 2,234 (1.01%) of the groups specified “other” as the race. We excluded these records leaving 218,348 for our analysis shown in Table 1. Note the empty cells in the age range up to range 24-35 had values from 0-9 but they were hidden in this public data set. We show these as “<10” in our tables.

**Table 1:** COVID death rates/100,000 from the NCHS monthly provisional death counts by age, sex, and race

|  |  | Black |  | Hispanic |  | Am. Indian<sup>a</sup> |  | Asian |  | White |  | All |  |
| --- | --- | --- | --- | --- | --- | --- | --- | --- | --- | --- | --- | --- | --- |
| Age | Sex | No. COVID deaths (deaths/100K) | No. in pop.<sup>b</sup> | No. COVID deaths (deaths/100K) | No. in pop. | No. COVID deaths (deaths/100K) | No. in pop. | No. COVID deaths (deaths/100K) | No. in pop. | No. COVID deaths (deaths/100K) | No. in pop. | No. COVID deaths (deaths/100K) | No. in pop. |
| 0-4 | All | -<sup>c</sup> (<10) | 2.99 M<sup>d</sup> | - (<10) | 5.09 M | - (<10) | 0.19 M | - (<10) | 1.13 M | - (<10) | 10.16 M | - (<10) | 19.58 M |
|  | F<sup>e</sup> | - (<10) | 1.47 M | - (<10) | 2.50 M | - (<10) | 0.10 M | - (<10) | 0.55 M | - (<10) | 4.95 M | - (<10) | 9.57 M |
|  | M<sup>f</sup> | - (<10) | 1.52 M | - (<10) | 2.60 M | - (<10) | 0.10 M | - (<10) | 0.58 M | - (<10) | 5.21 M | - (<10) | 10.01 M |
| 5-14 | All | - (<10) | 6.23 M | - (<10) | 10.57 M | - (<10) | 0.41 M | - (<10) | 2.42 M | - (<10) | 21.36 M | - (<10) | 40.99 M |
|  | F | - (<10) | 3.07 M | - (<10) | 5.18 M | - (<10) | 0.20 M | - (<10) | 1.19 M | - (<10) | 10.42 M | - (<10) | 20.05 M |
|  | M | - (<10) | 3.16 M | - (<10) | 5.39 M | - (<10) | 0.21 M | - (<10) | 1.24 M | - (<10) | 10.95 M | - (<10) | 20.94 M |
| 15-24 | All | 75 (1.2) | 6.42 M | 131 (1.3) | 9.88 M | - (<10) | 0.41 M | - (<10) | 2.67 M | 32 (<10) | 23.30 M | 238 (0.6) | 42.69 M |
|  | F | 35 (1.1) | 3.16 M | 32 (0.7) | 4.83 M | - (<10) | 0.20 M | - (<10) | 1.33 M | - (<10) | 11.35 M | 67 (<10) | 20.88 M |
|  | M | 40 (1.2) | 3.25 M | 99 (2.0) | 5.05 M | - (<10) | 0.21 M | - (<10) | 1.34 M | 32 (<10) | 11.95 M | 171 (0.8) | 21.81 M |
| 25-34 | All | 452 (6.6) | 6.81 M | 715 (7.5) | 9.53 M | 37 (8.7) | 0.43 M | 14 (<10) | 3.54 M | 293 (1.1) | 25.65 M | 1,511 (3.3) | 45.94 M |
|  | F | 183 (5.3) | 3.42 M | 214 (4.7) | 4.55 M | 12 (5.7) | 0.21 M | - (<10) | 1.80 M | 103 (0.8) | 12.60 M | 512 (2.3) | 22.58 M |
|  | M | 269 (7.9) | 3.38 M | 501 (10.1) | 4.98 M | 25 (11.6) | 0.21 M | 14 (0.8) | 1.74 M | 190 (1.5) | 13.04 M | 999 (4.3) | 23.36 M |
| 35-44 | All | 1,151 (21.0) | 5.48 M | 2,129 (24.6) | 8.66 M | 99 (29.3) | 0.34 M | 105 (3.2) | 3.29 M | 675 (2.8) | 23.90 M | 4,159 (10.0) | 41.66 M |
|  | F | 513 (17.8) | 2.88 M | 542 (12.9) | 4.20 M | 35 (20.4) | 0.17 M | 12 (0.7) | 1.74 M | 242 (2.0) | 11.88 M | 1,344 (6.4) | 20.87 M |
|  | M | 638 (24.5) | 2.60 M | 1,587 (35.6) | 4.46 M | 64 (38.5) | 0.17 M | 93 (6.0) | 1.55 M | 433 (3.6) | 12.01 M | 2,815 (13.5) | 20.79 M |
| 45-54 | All | 2,969 (57.0) | 5.21 M | 4,992 (70.0) | 7.13 M | 277 (86.7) | 0.32 M | 421 (15.0) | 2.80 M | 2,576 (10.1) | 25.41 M | 11,235 (27.5) | 40.87 M |
|  | F | 1,173 (42.3) | 2.77 M | 1,278 (36.1) | 3.54 M | 92 (55.7) | 0.17 M | 113 (7.6) | 1.49 M | 947 (7.4) | 12.73 M | 3,603 (17.4) | 20.70 M |

<sup>a</sup> American Indian
<sup>b</sup> No. in population calculated based on NCHS bridged race-ethnicity population estimates
<sup>c</sup> "-" (hyphen) indicates source data was suppressed for >10 deaths and estimates could not be computed
<sup>d</sup> Millions
<sup>e</sup> Female

|  |  |  |  |  |  |  |  |  |  |  |  |  |  |
| --- | --- | --- | --- | --- | --- | --- | --- | --- | --- | --- | --- | --- | --- |
|  | M | 1,796 (73.7) | 2.44 M | 3,714<br>(103.4) | 3.59 M | 185 (119.7) | 0.15 M | 308 (23.5) | 1.31 M | 1,629<br>(12.8) | 12.68 M | 7,632 (37.8) | 20.17 M |
| 55-64 | All | 7,505<br>(149.0) | 5.04 M | 9,000<br>(177.3) | 5.08 M | 490 (148.0) | 0.33 M | 1,142<br>(50.2) | 2.28 M | 9,239<br>(31.1) | 29.73 M | 27,376 (64.5) | 42.45 M |
|  | F | 3,152<br>(115.4) | 2.73 M | 2,695<br>(103.7) | 2.60 M | 209 (119.0) | 0.18 M | 311 (25.1) | 1.24 M | 3,402<br>(22.4) | 15.21 M | 9,769 (44.5) | 21.95 M |
|  | M | 4,353<br>(188.8) | 2.31 M | 6,305<br>(254.4) | 2.48 M | 281 (180.8) | 0.16 M | 831 (80.0) | 1.04 M | 5,837<br>(40.2) | 14.52 M | 17,607 (85.9) | 20.50 M |
| 65-74 | All | 11,569<br>(367.3) | 3.15 M | 11,156<br>(395.3) | 2.82 M | 602 (283.5) | 0.21 M | 1,995<br>(126.0) | 1.58 M | 21,787<br>(91.9) | 23.72 M | 47,109<br>(149.6) | 31.48 M |
|  | F | 4,841<br>(269.1) | 1.80 M | 3,961<br>(258.9) | 1.53 M | 275 (240.1) | 0.11 M | 690 (78.0) | 0.88 M | 8,596<br>(69.0) | 12.46 M | 18,363<br>(109.4) | 16.78 M |
|  | M | 6,728<br>(498.1) | 1.35 M | 7,195<br>(556.7) | 1.29 M | 327 (334.2) | 0.10 M | 1,305<br>(186.9) | 0.70 M | 13,191<br>(117.1) | 11.26 M | 28,746<br>(195.6) | 14.70 M |
| 75-84 | All | 10,891<br>(784.8) | 1.39 M | 9,887<br>(756.0) | 1.31 M | 487 (540.0) | 0.09 M | 2,187<br>(292.6) | 0.75 M | 35,008<br>(281.5) | 12.44 M | 58,460<br>(366.1) | 15.97 M |
|  | F | 5,267<br>(619.6) | 0.85 M | 4,246<br>(557.4) | 0.76 M | 237 (469.2) | 0.05 M | 853 (203.1) | 0.42 M | 15,703<br>(227.9) | 6.89 M | 26,306<br>(293.2) | 8.97 M |
|  | M | 5,624<br>(1,046.0) | 0.54 M | 5,641<br>(1,033.1) | 0.55 M | 250 (630.2) | 0.04 M | 1,334<br>(407.4) | 0.33 M | 19,305<br>(348.0) | 5.55 M | 32,154<br>(459.5) | 7.00 M |
| 85+ | All | 8,357<br>(1,609.2) | 0.52 M | 7,611<br>(1,495.0) | 0.51 M | 280 (927.0) | 0.03 M | 2,567<br>(841.8) | 0.30 M | 49,445<br>(943.4) | 5.24 M | 68,260<br>(1,033.5) | 6.60 M |
|  | F | 5,261<br>(1,458.1) | 0.36 M | 4,116<br>(1,271.3) | 0.32 M | 155 (813.7) | 0.02 M | 1,509<br>(792.0) | 0.19 M | 29,820<br>(894.3) | 3.33 M | 40,861<br>(966.3) | 4.23 M |
|  | M | 3,096<br>(1,953.3) | 0.16 M | 3,495<br>(1,885.9) | 0.19 M | 125<br>(1120.4) | 0.01 M | 1,058<br>(924.8) | 0.11 M | 19,625<br>(1,029.0) | 1.91 M | 27,399<br>(1,152.9) | 2.38 M |
| All |  | 42,969<br>(99.4) | 43.24 M | 45,621<br>(75.3) | 60.57<br>M | 2,272 (82.2) | 2.77 M | 8,431<br>(40.6) | 20.76 M | 119,055<br>(59.3) | 200.91 M | 218,348<br>(66.5) | 328.24<br>M |

Ignoring age and sex, overall death rates were highest at 99 deaths/100,000 for Blacks, and they decrease in order to 82 for American Indians, 75, for Hispanics, 59 for Whites, and 41 for Asians, per 100,000. Overall, deaths rated climbed almost geometrically with age (see Table 1), so much so, that seventy percent of all COVID-19 deaths occurred in individual over age 70. The highest COVID-19 death rates occurred in males over age 85. Among such Black patients, it was 1,953, Hispanics 1,886, American Indians 1,120, Whites 1,029 and Asians 925, deaths per 100,000. In every age/race category, the male COVID-19 death rate exceeded that of the females.

The difference in the COVID-19 death prevalence in all 10 pairwise comparisons between racial categories was significant at p < 0.001 after Bonferroni’s correction for multiple comparisons. We used a more recent set of death records from weeks 44 through 47 (10/21/2020 to 11/21/2020) which did not yet have race information, to assess the death rate during the current surge in COVID-19 cases (Table 2).^16^ Among elders over age 74, 22,000 COVID-19 deaths occurred over four weeks of this surge and that death rate is expected likely to continue, adding urgency to protecting this age group.

**Table 2:** Provisional COVID-19 Death Counts by Sex and Age during recent surge (Weeks 44-47). No race/ethnicity data yet available.

|  | Week 44 (ending 10/21/2020) |  | Week 45 (ending 11/7/2020) |  | Week 46 (ending 11/7/2020) |  | Week 47 (ending 11/21/2020) |  |  |  |
| --- | --- | --- | --- | --- | --- | --- | --- | --- | --- | --- |
| Age | No. cum. Deaths (%) | No. deaths (%) | No. cum. Deaths (%) | No. deaths (%) | No. cum. Deaths (%) | No. deaths (%) | No. cum. Deaths (%) | No. deaths (%) | Total COVID deaths (Weeks 44-47) | Cum total COVID deaths (Weeks 44-47) |
| Under 1 | 6,289 (100%)<sup>g</sup> | 0 (0%) | 7,572 (100%) | 1 (0%) | 8,682 (100%) | 0 (0%) | 8,955 (100%) | 0 (0%) | 1 | 31,498 |
| Over 1 | 6,289 (200%) | 0 (0%) | 7,571 (200%) | 0 (0%) | 8,682 (200%) | 0 (0%) | 8,955 (200%) | 1 (0%) | 1 | 31,497 |
| Over 5 | 6,289 (200%) | 2 (0%) | 7,571 (200%) | 1 (0%) | 8,682 (200%) | 1 (0%) | 8,954 (200%) | 1 (0%) | 5 | 31,496 |
| Over 15 | 6,287 (200%) | 12 (0%) | 7,570 (200%) | 8 (0%) | 8,681 (200%) | 5 (0%) | 8,953 (200%) | 9 (0%) | 34 | 31,491 |
| Over 25 | 6,275 (200%) | 32 (1%) | 7,562 (200%) | 42 (1%) | 8,676 (200%) | 36 (0%) | 8,944 (200%) | 38 (0%) | 148 | 31,457 |
| Over 35 | 6,243 (199%) | 97 (2%) | 7,520 (199%) | 94 (1%) | 8,640 (200%) | 88 (1%) | 8,906 (199%) | 92 (1%) | 371 | 31,309 |
| Over 45 | 6,146 (198%) | 237 (4%) | 7,426 (198%) | 269 (4%) | 8,552 (199%) | 311 (4%) | 8,814 (198%) | 268 (3%) | 1,085 | 30,938 |
| Over 55 | 5,909 (194%) | 649 (10%) | 7,157 (195%) | 718 (9%) | 8,241 (195%) | 838 (10%) | 8,546 (195%) | 834 (9%) | 3,039 | 29,853 |
| Over 65 | 5,260 (184%) | 1,298 (21%) | 6,439 (185%) | 1,566 (21%) | 7,403 (185%) | 1,734 (20%) | 7,712 (186%) | 1,758 (20%) | 6,356 | 26,814 |
| Over 75 | 3,962 (163%) | 1,839 (29%) | 4,873 (164%) | 2,270 (30%) | 5,669 (165%) | 2,645 (30%) | 5,954 (166%) | 2,665 (30%) | 9,419 | 20,458 |
| Over 85 | 2,123 (134%) | 2,123 (34%) | 2,603 (134%) | 2,603 (34%) | 3,024 (135%) | 3,024 (35%) | 3,289 (137%) | 3,289 (37%) | 11,039 | 11,039 |
| All Ages |  | 6,289 (100%) |  | 7,572 (100%) |  | 8,682 (100%) |  | 8,955 (100%) | 31,498 |  |

### Breakdown of case rates

The CDC case surveillance data set^6^ contained 8,242,333 COVID-19 cases from April 1, when an ICD code for COVID-19 was published, until October 30, the date of the most recent data. Age and sex data were missing in trivial amounts (0.14%) and (1.17%). However, definitive race status was absent from 43.6% of these. We used the 4,646,942 records with complete age, sex, and race data to report case rates, broken down by age, sex, and race (Supplemental Table 1).

Ignoring age and sex, American Indians had the highest case rates at 2,133 per 100,000. The rate for Hispanics, Blacks, Whites and Asians followed in decreasing order of 1935, 1655, 1267 and 744, per 100,000, respectively. The Asian case rate was one third that of that of American Indians.

Among Blacks, the case rates peak in the 80+ age range, among Hispanics in the 40-49 age range, and among Whites and Asians, in the 20-29-year age range. American Indian case rates peaked in the 30-39 age range to the highest level of any age/race category.

Among American Indians, the transition from more female to more male cases occurred at age 80; among Blacks, at age 60; and among Hispanics and Whites, at age 50. Among Asians the sexes had similar case rates at all ages, and the sex with the most cases varied with age range. Ignoring race, the highest case prevalence of any age/sex category (2167/100,000) occurred among 20-29-year-old women, a rate almost 20% greater than the next closest age/sex category.

The difference in the COVID-19 case prevalence in all 10 pairwise comparisons between racial categories was significant at p < 0.001 after Bonferroni’s correction for multiple comparisons.

### Logistic regression model predicting COVID-19 deaths and COVID-19 case

Table 3 presents the logistic regression estimate for likelihood of a COVID-19 death for each demographic in order from highest to lowest estimated risk. Eighty-five plus year old Black and Hispanic men (see rows 1 and 2) have the worst risk. Columns A-C carry the values of race, age and sex that define each category. Column D carries the number of COVID-19 deaths and Column E the number of individuals in each category. Column F carries logistic estimated death risk and its 95% confidence intervals for each category. We expressed these risk in terms of number per 100,000, rather than pure probabilities to fit public health reporting conventions. In a given row, Columns G and H carry are the cumulative sum of all of the deaths, and number of people, respectively in all of the categories at or above that row. One can think of the cumulative sum of people (column H) to be the number of people to vaccinate in order to prevent the number of deaths in Column G.

**Table 3:** Logistic regression on NCHS COVID death data showing the risk of COVID-19 death for each age/race/sex risk group sorted in order by death risk/100,000 U.S. inhabitants

|  | A | B | C | D | E | F | I | J |
| --- | --- | --- | --- | --- | --- | --- | --- | --- |
|  | Race | Age | Sex | No. COVID Death | No. in Pop. | Death risk /100,000 (95% CI) | Cum. No. of deaths | Cum. No. in population |
| 1 | Black | 85+ | Male | 3,096 | 0.16 M | 2,160.7 (2,114.3 - 2,208.2) | 3,096 | 0.16 M |

<sup>g</sup> Cumulation order is the reverse of the usual. It provides the counts and percentage of COVID deaths at or below the current row

|  |  |  |  |  |  |  |  |  |
| --- | --- | --- | --- | --- | --- | --- | --- | --- |
| 2 | Hispanic | 85+ | Male | 3,495 | 0.19 M | 1,951.6 (1,907.9 - 1,996.2) | 6,591 | 0.34 M |
| 3 | Black | 85+ | Female | 5,261 | 0.36 M | 1,367.0 (1,337.8 - 1,396.8) | 11,852 | 0.70 M |
| 4 | Hispanic | 85+ | Female | 4,116 | 0.32 M | 1,233.7 (1,206.0 - 1,262.0) | 15,968 | 1.03 M |
| 5 | White | 85+ | Male | 19,625 | 1.91 M | 1,233.3 (1,221.3 - 1,245.4) | 35,593 | 2.94 M |
| 6 | Am. Indian<sup>h</sup> | 85+ | Male | 125 | 0.01 M | 1,208.7 (1,075.9 - 1,357.8) | 35,718 | 2.95 M |
| 7 | Asian | 85+ | Male | 1,058 | 0.11 M | 1,094.8 (1,053.3 - 1,138.0) | 36,776 | 3.06 M |
| 8 | Black | 75-84 | Male | 5,624 | 0.54 M | 1,014.9 (995.6 - 1,034.5) | 42,400 | 3.60 M |
| 9 | Hispanic | 75-84 | Male | 5,641 | 0.55 M | 963.9 (944.8 - 983.3) | 48,041 | 4.14 M |
| 10 | White | 85+ | Female | 29,820 | 3.33 M | 777.5 (770.1 - 785.1) | 77,861 | 7.48 M |
| 11 | Am. Indian | 85+ | Female | 155 | 0.02 M | 762.0 (677.9 - 856.4) | 78,016 | 7.50 M |
| 12 | Asian | 85+ | Female | 1,509 | 0.19 M | 689.9 (663.6 - 717.2) | 79,525 | 7.69 M |
| 13 | Am. Indian | 75-84 | Male | 250 | 0.04 M | 681.6 (623.8 - 744.7) | 79,775 | 7.73 M |
| 14 | Black | 75-84 | Female | 5,267 | 0.85 M | 639.3 (627.1 - 651.7) | 85,042 | 8.58 M |
| 15 | Hispanic | 75-84 | Female | 4,246 | 0.76 M | 607.0 (594.9 - 619.4) | 89,288 | 9.34 M |
| 16 | Hispanic | 65-74 | Male | 7,195 | 1.29 M | 494.9 (485.7 - 504.3) | 96,483 | 10.63 M |
| 17 | Black | 65-74 | Male | 6,728 | 1.35 M | 466.2 (457.6 - 474.9) | 103,211 | 11.98 M |
| 18 | Am. Indian | 75-84 | Female | 237 | 0.05 M | 428.8 (392.4 - 468.7) | 103,448 | 12.03 M |
| 19 | Asian | 75-84 | Male | 1,334 | 0.33 M | 369.8 (354.6 - 385.7) | 104,782 | 12.36 M |
| 20 | Am. Indian | 65-74 | Male | 327 | 0.10 M | 354.5 (327.3 - 383.9) | 105,109 | 12.46 M |
| 21 | White | 75-84 | Male | 19,305 | 5.55 M | 354.5 (350.5 - 358.4) | 124,414 | 18.01 M |
| 22 | Hispanic | 65-74 | Female | 3,961 | 1.53 M | 311.2 (305.2 - 317.2) | 128,375 | 19.54 M |
| 23 | Black | 65-74 | Female | 4,841 | 1.80 M | 293.1 (287.6 - 298.6) | 133,216 | 21.34 M |
| 24 | Asian | 75-84 | Female | 853 | 0.42 M | 232.4 (222.8 - 242.4) | 134,069 | 21.76 M |
| 25 | Am. Indian | 65-74 | Female | 275 | 0.11 M | 222.8 (205.6 - 241.3) | 134,344 | 21.87 M |
| 26 | White | 75-84 | Female | 15,703 | 6.89 M | 222.7 (220.2 - 225.3) | 150,047 | 28.76 M |
| 27 | Hispanic | 55-64 | Male | 6,305 | 2.48 M | 219.0 (214.4 - 223.6) | 156,352 | 31.24 M |
| 28 | Black | 55-64 | Male | 4,353 | 2.31 M | 186.7 (182.5 - 191.0) | 160,705 | 33.54 M |
| 29 | Am. Indian | 55-64 | Male | 281 | 0.16 M | 184.4 (168.8 - 201.5) | 160,986 | 33.70 M |
| 30 | Asian | 65-74 | Male | 1,305 | 0.70 M | 159.1 (152.3 - 166.3) | 162,291 | 34.40 M |

<sup>h</sup> American Indian

|  |  |  |  |  |  |  |  |  |
| --- | --- | --- | --- | --- | --- | --- | --- | --- |
| 31 | Hispanic | 55-64 | Female | 2,695 | 2.60 M | 137.5 (134.6 - 140.5) | 164,986 | 36.99 M |
| 32 | Black | 55-64 | Female | 3,152 | 2.73 M | 117.2 (114.6 - 120.0) | 168,138 | 39.73 M |
| 33 | Am. Indian | 55-64 | Female | 209 | 0.18 M | 115.8 (106.0 - 126.5) | 168,347 | 39.90 M |
| 34 | White | 65-74 | Male | 13,191 | 11.26 M | 114.2 (112.6 - 115.8) | 181,538 | 51.16 M |
| 35 | Am. Indian | 45-54 | Male | 185 | 0.15 M | 107.3 (95.4 - 120.7) | 181,723 | 51.32 M |
| 36 | Asian | 65-74 | Female | 690 | 0.88 M | 99.9 (95.6 - 104.4) | 182,413 | 52.20 M |
| 37 | Hispanic | 45-54 | Male | 3,714 | 3.59 M | 85.9 (83.5 - 88.3) | 186,127 | 55.79 M |
| 38 | White | 65-74 | Female | 8,596 | 12.46 M | 71.7 (70.7 - 72.7) | 194,723 | 68.25 M |
| 39 | Black | 45-54 | Male | 1,796 | 2.44 M | 71.0 (68.5 - 73.6) | 196,519 | 70.69 M |
| 40 | Am. Indian | 45-54 | Female | 92 | 0.17 M | 67.4 (59.9 - 75.8) | 196,611 | 70.85 M |
| 41 | Asian | 55-64 | Male | 831 | 1.04 M | 62.9 (59.4 - 66.7) | 197,442 | 71.89 M |
| 42 | Hispanic | 45-54 | Female | 1,278 | 3.54 M | 53.9 (52.4 - 55.5) | 198,720 | 75.43 M |
| 43 | Black | 45-54 | Female | 1,173 | 2.77 M | 44.6 (43.0 - 46.2) | 199,893 | 78.20 M |
| 44 | Asian | 55-64 | Female | 311 | 1.24 M | 39.5 (37.3 - 41.9) | 200,204 | 79.44 M |
| 45 | White | 55-64 | Male | 5,837 | 14.52 M | 38.4 (37.6 - 39.2) | 206,041 | 93.96 M |
| 46 | Am. Indian | 35-44 | Male | 64 | 0.17 M | 36.1 (29.7 - 44.0) | 206,105 | 94.13 M |
| 47 | Hispanic | 35-44 | Male | 1,587 | 4.46 M | 30.0 (28.8 - 31.3) | 207,692 | 98.59 M |
| 48 | Black | 35-44 | Male | 638 | 2.60 M | 26.1 (24.6 - 27.7) | 208,330 | 101.19 M |
| 49 | White | 55-64 | Female | 3,402 | 15.21 M | 24.1 (23.6 - 24.6) | 211,732 | 116.40 M |
| 50 | Am. Indian | 35-44 | Female | 35 | 0.17 M | 22.7 (18.6 - 27.6) | 211,767 | 116.57 M |
| 51 | Hispanic | 35-44 | Female | 542 | 4.20 M | 18.8 (18.0 - 19.7) | 212,309 | 120.76 M |
| 52 | Asian | 45-54 | Male | 308 | 1.31 M | 18.7 (17.0 - 20.6) | 212,617 | 122.07 M |
| 53 | Black | 35-44 | Female | 513 | 2.88 M | 16.4 (15.5 - 17.4) | 213,130 | 124.95 M |
| 54 | White | 45-54 | Male | 1,629 | 12.68 M | 12.5 (12.0 - 13.0) | 214,759 | 137.63 M |
| 55 | Asian | 45-54 | Female | 113 | 1.49 M | 11.8 (10.7 - 12.9) | 214,872 | 139.12 M |
| 56 | Am. Indian | 25-34 | Male | 25 | 0.21 M | 10.7 (7.7 - 14.7) | 214,897 | 139.34 M |
| 57 | Hispanic | 25-34 | Male | 501 | 4.98 M | 9.1 (8.5 - 9.8) | 215,398 | 144.32 M |
| 58 | Black | 25-34 | Male | 269 | 3.38 M | 8.2 (7.5 - 9.0) | 215,667 | 147.70 M |
| 59 | White | 45-54 | Female | 947 | 12.73 M | 7.8 (7.5 - 8.1) | 216,614 | 160.43 M |
| 60 | Am. Indian | 25-34 | Female | 12 | 0.21 M | 6.7 (4.8 - 9.2) | 216,626 | 160.65 M |
| 61 | Hispanic | 25-34 | Female | 214 | 4.55 M | 5.7 (5.3 - 6.2) | 216,840 | 165.19 M |
| 62 | Black | 25-34 | Female | 183 | 3.42 M | 5.1 (4.7 - 5.6) | 217,023 | 168.61 M |
| 63 | Asian | 35-44 | Male | 93 | 1.55 M | 4.0 (3.3 - 4.8) | 217,116 | 170.16 M |
| 64 | White | 35-44 | Male | 433 | 12.01 M | 3.5 (3.2 - 3.7) | 217,549 | 182.18 M |
| 65 | Asian | 35-44 | Female | 12 | 1.74 M | 2.5 (2.1 - 3.0) | 217,561 | 183.92 M |
| 66 | White | 35-44 | Female | 242 | 11.88 M | 2.2 (2.0 - 2.3) | 217,803 | 195.80 M |
| 67 | Hispanic | 15-24 | Male | 99 | 5.05 M | 1.6 (1.4 - 1.9) | 217,902 | 200.85 M |
| 68 | Black | 15-24 | Male | 40 | 3.25 M | 1.4 (1.1 - 1.8) | 217,942 | 204.10 M |
| 69 | White | 25-34 | Male | 190 | 13.04 M | 1.4 (1.2 - 1.6) | 218,132 | 217.15 M |
| 70 | Hispanic | 15-24 | Female | 32 | 4.83 M | 1.0 (0.9 - 1.2) | 218,164 | 221.98 M |
| 71 | Black | 15-24 | Female | 35 | 3.16 M | 0.9 (0.7 - 1.1) | 218,199 | 225.14 M |
| 72 | White | 25-34 | Female | 103 | 12.60 M | 0.9 (0.8 - 1.0) | 218,302 | 237.75 M |
| 73 | Asian | 25-34 | Male | 14 | 1.74 M | 0.5 (0.3 - 0.8) | 218,316 | 239.48 M |
| 74 | Asian | 25-34 | Female | * | 1.80 M | 0.3 (0.2 - 0.5) | 218,316 | 241.28 M |
| 75 | White | 15-24 | Male | 32 | 11.95 M | 0.2 (0.1 - 0.2) | 218,348 | 253.23 M |
| 76 | White | 15-24 | Female | * | 11.35 M | 0.1 (0.1 - 0.1) | 218,348 | 264.58 M |
| 77 | Black | 0-4 | Male | * | 1.52 M | † | † | 266.11 M |
| 78 | Am. Indian | 0-4 | Male | * | 0.10 M | † | † | 266.20 M |
| 79 | Hispanic | 0-4 | Male | * | 2.60 M | † | † | 268.80 M |
| 80 | White | 0-4 | Male | * | 5.21 M | † | † | 274.01 M |
| 81 | Asian | 0-4 | Male | * | 0.58 M | † | † | 274.59 M |
| 82 | White | 5-14 | Male | * | 10.95 M | † | † | 285.54 M |
| 83 | Black | 5-14 | Male | * | 3.16 M | † | † | 288.70 M |
| 84 | Am. Indian | 15-24 | Male | * | 0.21 M | † | † | 288.91 M |
| 85 | Am. Indian | 5-14 | Male | * | 0.21 M | † | † | 289.12 M |
| 86 | Hispanic | 5-14 | Male | * | 5.39 M | † | † | 294.51 M |
| 87 | Asian | 15-24 | Male | * | 1.34 M | † | † | 295.85 M |
| 88 | Asian | 5-14 | Male | * | 1.24 M | † | † | 297.09 M |
| 89 | Black | 0-4 | Female | * | 1.47 M | † | † | 298.56 M |
| 90 | Am. Indian | 0-4 | Female | * | 0.10 M | † | † | 298.66 M |
| 91 | Hispanic | 0-4 | Female | * | 2.50 M | † | † | 301.15 M |
| 92 | White | 0-4 | Female | * | 4.95 M | † | † | 306.11 M |
| 93 | Asian | 0-4 | Female | * | 0.55 M | † | † | 306.66 M |
| 94 | White | 5-14 | Female | * | 10.42 M | † | † | 317.07 M |
| 95 | Black | 5-14 | Female | * | 3.07 M | † | † | 320.14 M |
| 96 | Am. Indian | 15-24 | Female | * | 0.20 M | † | † | 320.34 M |
| 97 | Am. Indian | 5-14 | Female | * | 0.20 M | † | † | 320.55 M |
| 98 | Hispanic | 5-14 | Female | * | 5.18 M | † | † | 325.73 M |
| 99 | Asian | 15-24 | Female | * | 1.33 M | † | † | 327.05 M |
| 100 | Asian | 5-14 | Female | * | 1.19 M | † | † | 328.24 M |

\* Source data was suppressed for >10 deaths
† Could not estimate because of suppressed data

For a given sized vaccine supply one could choose the allocation of vaccine to risk groups by reading down from the first row in Table 3 until the cumulative number of individuals (Column H) first exceeds the vaccine supply, and then vaccinate all people in risk groups above that row, and a portion of the people in that row as needed consume the whole supply. For example, if one had enough supply for 12 million individuals (2 shots per person), one could vaccinate all risk groups from row 1 to 17 (highlighted with arrow on Table 3, and 200,000 in the category of row 18 to finish off the supply. Doing so would consume only 3.7% of the vaccine supply required for the whole U.S. population and prevent 47% of all COVID deaths, a highly leveraged use.

Supplemental Table 2 shows the results of an analogous logistic analysis of cases (infections) based on 4,646,942 records with complete age/sex/race data, with COVID-19 cases as outcomes. Female American Indians aged 30-39 have the highest risk of becoming infected with COVID-19 (becoming a case). Most of the top risk categories are occupied by American Indian or Hispanic females in different age ranges. If we dedicated the a vaccine supply of 14 million to individuals with the highest infection risk rather than the highest death risk, we would vaccinate all of the demographic categories in supplemental Table 2 from row 1 to row 7 (highlighted by an arrow in supplemental table 2), which would include 5.7% of the total population and would only prevent 11% of the COVID-19 cases. The results for our analysis of the 8,120,644, with complete age and sex data (not shown) were similar. Vaccinating 6.7% of the individuals with highest case risk would prevent only 10% of cases. In both cases, the payoff for vaccinating based on case risk was a meager one fifth of the payoff based on death risk.

### Cases and deaths among special populations

Any allocation rule for SARS-CoV-2 vaccine will want to consider the infection and death rates in some special population for prioritization. Among nursing home residents, the cumulative number of deaths per 100,000 is staggering at 5,188/100,000. (See Table 4, row 1).

**Table 4:** Deaths and population sized of COVID special risk categories

| Special risk category | Number of COVID-19 deaths | Number in the category | # Deaths/100,000 |
| --- | --- | --- | --- |
| Nursing home (long term care) | 72,642<sup>8</sup> | 1,400,000<sup>22</sup> | 5,188 |
| Prison | 1,494<sup>13</sup> | 2,300,000<sup>12</sup> | 64.9 |
| Health Care Workers | 1,700<sup>10</sup><br>847<sup>11</sup> | 16,866,020<sup>17</sup><br>20,000,000<sup>18</sup> | 10 |

Data collected from the Department of Corrections websites are principally represented by prisons, with exception of a few states that include jails.^13^ For prisoners, the death rate was a modest 64.9/100,000 -- about the same COVID-19 death risk as the average U.S. 55-65-year-olds (Table 1) and by the numbers would be prioritized after the 45 highest risk categories in Table 3 (arrow highlights row 45 in table 3)..

Data about COVID-19 deaths cases among healthcare workers (HCWs) are limited. However, if we consider all 14 to 20 million HCWs^17,18^ to be at equal risk of COVID-19 death, their average death risk is 10/100,000 (Table 4), which is less than the 12/100,000 risk of death in an auto accident^19^ in the United States. This low figure suggests the denominator includes a large number of HCWs with a very low risk of COVID-19 death, which dilute the real COVID death risk of a smaller number of frontline HCWs.

## DISCUSSION

We created two simple statistical models using logistic regression, one for predicting COVID-19 deaths and another for predicting COVID-19 cases. Babus, Das, and Lee^20^ report a more complicated model but only considered infections (cases) as outcome.

We used NCHS’s vetted COVID-19 death records using only age ranges, race and sex as predictors of COVID-19 deaths. Our model successfully distinguished death risk by demographic category, over a two orders of magnitude range of risks. However, it would probably produce even better estimates if more data were available, such as, actual ages (which are available to some agencies), rather than 10-year ranges, and case, or death, rates by geographic location for targeting available vaccine supplies. New York State plans to factor location risk by ranking high risk locations over low ones^21^ but location risk could be included in a formal logistic model directly.

Demographic breakdowns were not available for special risk groups, so we generated simple overall risk figures based on available death and population counts (Table 4). The risk of COVID deaths among the 1.3 million nursing home residents^22^ is extreme at 5,188 per 100,000; which puts them at a level well above that of 85+ year old black males and in front of the line for SARS-CoV-2 vaccination. The comparable risk for 2.3 million prison residents at 64.9/100,000, is slightly above the overall death rate of 55-64-year-olds at 64.9 (Table 1) and below that of 41 demographic groups with the highest COVID-19 death risk.

Front line HCWs deserve special priority because they brave COVID-19 hot zones. However, a large share of the nominal 14-20 million HCWs are not front line. The risks of administrators and pharmacist preparing intravenous fluids in the hospital basement are exponentially less than a nurse anesthetist or emergency room physician examining or intubating COVID-19 patients. Careful discrimination between the levels of HCW risks based on degree of COVID-19 patient contact will be important to ensure that the highest risk HCWs are vaccinated first and that some of the early supplies go to the high-risk elderly rather than minimum risk HCWs. A few states are distinguishing HCWs by risk level. New York called out ICU and emergency room HCWs for special priority.^21^

Some states propose to allocate vaccine supply by occupation class beyond HCWs.^21^ For example, New York State’s vaccination program places some essential workers, e.g. teachers, grocery workers, pharmacists, transit workers, in line for vaccination ahead of their senior citizens (age 65 and above).^22^ Some states put all essential workers who cannot work from home (WFH) ahead of their elderly.^23^ However, the early vaccine supplies will not come close to accommodating all such essential workers, whose numbers range from 55 to 80 million and whose ages ranges predict a lowish COVID death risk.^24,25^ However, early supplies will not be enough for the 54 million U.S elders who are over 65 either. It will take a narrower focus on a smaller number of the highest risk individuals to fit early supplies. An example would be the 12 million individuals we described in the results section which would not require an exorbitant supply but could prevent 47% of COVID deaths. Relevant to racial equity, 65% of that 12 million chosen on the basis of their COVID-19 death risk would be minorities.

Importantly, 75+ year old U.S. seniors are dying from COVID-19 at rate of 20,000 lives per month which rate could be reduced by timely SARS-CoV-2 vaccination. Recognizing the time pressure, Great Britain has committed an early tranche of their vaccine supply to its 80+ year old citizens.^26^

We hope that the statistical ranking method and the risk data it produced will be of use to state governments now working on plans for allocating their SARS-CoV-2 vaccine supplies.

## Data Availability

All data used in the article are publicly available from the relevant citation included in the text. Code for the analysis may be obtained from the authors upon request.

## Acknowledgements

This research was supported by the Intramural Research Program of the National Library of Medicine, National Institutes of Health.

CJM had full access to all the data in the study and takes responsibility for the integrity of the data and the accuracy of the data analysis.

