## Supplemental Tables for "A method for prioritizing risk groups for early SARS-CoV-2 Vaccination, By the Numbers"

Table 1 COVID-19 Case rates/100,000 from CDC COVID-19 public surveillance data broken down by age, sex and race

|  |  | Black | | Hispanic | | Am. Indian^[1]^ | | Asian | | White | | All | |
| --- | --- | --- | --- | --- | --- | --- | --- | --- | --- | --- | --- | --- | --- |
| Age | Sex | No. COVID cases (cases/  100K) | No. in pop.^[2]^ | No. COVID cases (cases/  100K) | No. in pop. | No. COVID cases (cases/  100K) | No. in pop. | No. COVID cases (cases/  100K) | No. in pop. | No. COVID cases (cases/  100K) | No. in pop. | No. COVID cases (cases/  100K) | No. in pop. |
| 0 - 9 | All | 27,823 (458) | 6.10 M^[3]^ | 63,847 (618) | 10.00 M | 3,577 (898) | 0.40 M | 6,337 (273) | 2.30 M | 70,292 (341) | 20.60 M | 171,876 (432) | 39.80 M |
|  | F^[4]^ | 13,654 (457) | 3.00 M | 31,107 (614) | 5.10 M | 1,827 (934) | 0.20 M | 2,927 (259) | 1.10 M | 33,967 (338) | 10.10 M | 83,482 (429) | 19.40 M |
|  | M^[5]^ | 14,169 (459) | 3.10 M | 32,740 (621) | 5.30 M | 1,750 (864) | 0.20 M | 3,410 (286) | 1.20 M | 36,325 (343) | 10.60 M | 88,394 (435) | 20.30 M |
| 10 - 19 | All | 58,985 (938) | 6.30 M | 133,145 (1,285) | 10.40 M | 7,141 (1,738) | 0.40 M | 13,063 (524) | 2.50 M | 258,917 (1,161) | 22.30 M | 471,251 (1,126) | 41.90 M |
|  | F | 30,480 (983) | 3.10 M | 68,694 (1,352) | 5.10 M | 3,735 (1,841) | 0.20 M | 6,504 (526) | 1.20 M | 137,951 (1,269) | 10.90 M | 247,364 (1,207) | 20.50 M |
|  | M | 28,505 (893) | 3.20 M | 64,451 (1,220) | 5.30 M | 3,406 (1,637) | 0.20 M | 6,559 (522) | 1.30 M | 120,966 (1,059) | 11.40 M | 223,887 (1,048) | 21.40 M |
| 20 - 29 | All | 129,865 (1,871) | 6.90 M | 242,031 (2,474) | 9.80 M | 10,979 (2,505) | 0.40 M | 31,550 (1,006) | 3.10 M | 469,614 (1,890) | 24.80 M | 884,039 (1,958) | 45.10 M |
|  | F | 74,736 (2,177) | 3.40 M | 127,038 (2,684) | 4.70 M | 6,085 (2,814) | 0.20 M | 16,083 (1,028) | 1.60 M | 254,471 (2,099) | 12.10 M | 478,413 (2,167) | 22.10 M |
|  | M | 55,129 (1,572) | 3.50 M | 114,993 (2,277) | 5.10 M | 4,894 (2,203) | 0.20 M | 15,467 (984) | 1.60 M | 215,143 (1,692) | 12.70 M | 405,626 (1,758) | 23.10 M |
| 30 - 39 | All | 126,168 (2,097) | 6.00 M | 227,740 (2,504) | 9.10 M | 10,873 (2,895) | 0.40 M | 29,814 (843) | 3.50 M | 344,793 (1,371) | 25.10 M | 739,388 (1,674) | 44.20 M |
|  | F | 72,451 (2,334) | 3.10 M | 114,899 (2,647) | 4.30 M | 5,933 (3,141) | 0.20 M | 15,356 (834) | 1.80 M | 183,972 (1,477) | 12.50 M | 392,611 (1,790) | 21.90 M |
|  | M | 53,717 (1,844) | 2.90 M | 112,841 (2,373) | 4.80 M | 4,940 (2,647) | 0.20 M | 14,458 (852) | 1.70 M | 160,821 (1,268) | 12.70 M | 346,777 (1,559) | 22.20 M |
| 40 - 49 | All | 109,787 (2,098) | 5.20 M | 212,354 (2,667) | 8.00 M | 8,861 (2,789) | 0.30 M | 24,492 (799) | 3.10 M | 332,223 (1,399) | 23.70 M | 687,717 (1,706) | 40.30 M |
|  | F | 62,765 (2,261) | 2.80 M | 107,877 (2,750) | 3.90 M | 4,735 (2,912) | 0.20 M | 13,098 (802) | 1.60 M | 176,915 (1,495) | 11.80 M | 365,390 (1,798) | 20.30 M |
|  | M | 47,022 (1,914) | 2.50 M | 104,477 (2,586) | 4.00 M | 4,126 (2,660) | 0.20 M | 11,394 (795) | 1.40 M | 155,308 (1,304) | 11.90 M | 322,327 (1,612) | 20.00 M |
| 50 - 59 | All | 109,450 (2,087) | 5.20 M | 155,092 (2,517) | 6.10 M | 7,997 (2,386) | 0.30 M | 21,664 (870) | 2.50 M | 378,200 (1,345) | 28.10 M | 672,403 (1,588) | 42.40 M |
|  | F | 60,604 (2,160) | 2.80 M | 77,743 (2,521) | 3.10 M | 4,326 (2,471) | 0.20 M | 11,349 (850) | 1.30 M | 195,134 (1,372) | 14.20 M | 349,156 (1,615) | 21.60 M |
|  | M | 48,846 (2,004) | 2.40 M | 77,349 (2,514) | 3.10 M | 3,671 (2,292) | 0.20 M | 10,315 (892) | 1.20 M | 183,066 (1,317) | 13.90 M | 323,247 (1,559) | 20.70 M |
| 60 - 69 | All | 82,249 (1,948) | 4.20 M | 80,599 (2,083) | 3.90 M | 5,524 (1,978) | 0.28 M | 14,980 (758) | 2.00 M | 306,946 (1,109) | 27.70 M | 490,298 (1,289) | 38.00 M |
|  | F | 44,100 (1,881) | 2.30 M | 39,904 (1,959) | 2.00 M | 3,021 (2,011) | 0.15 M | 7,713 (706) | 1.10 M | 153,478 (1,070) | 14.30 M | 248,216 (1,243) | 20.00 M |
|  | M | 38,149 (2,032) | 1.90 M | 40,695 (2,222) | 1.80 M | 2,503 (1,939) | 0.13 M | 7,267 (823) | 0.90 M | 153,468 (1,151) | 13.30 M | 242,082 (1,341) | 18.10 M |
| 70 - 79 | All | 43,309 (2,014) | 2.10 M | 35,688 (1,819) | 2.00 M | 2,691 (1,842) | 0.15 M | 7,191 (636) | 1.10 M | 204,159 (1,116) | 18.20 M | 293,038 (1,237) | 23.70 M |
|  | F | 23,241 (1,838) | 2.30 M | 17,947 (1,633) | 1.10 M | 1,520 (1,916) | 0.08 M | 3,619 (573) | 0.60 M | 103,470 (1,057) | 9.80 M | 149,797 (1,165) | 12.90 M |
|  | M | 20,068 (2,264) | 0.90 M | 17,741 (2,056) | 0.90 M | 1,171 (1,753) | 0.07 M | 3,572 (715) | 0.50 M | 100,689 (1,184) | 8.50 M | 143,241 (1,324) | 10.80 M |
| 80+ | All | 27,851 (2,622) | 1.10 M | 21,285 (2,053) | 1.00 M | 1,355 (2,115) | 0.06 M | 5,426 (900) | 0.60 M | 181,015 (1,782) | 10.10 M | 236,932 (1,834) | 12.90 M |
|  | F | 17,529 (2,495) | 0.70 M | 12,433 (1,953) | 0.60 M | 841 (2,179) | 0.04 M | 3,170 (881) | 0.40 M | 112,207 (1,831) | 6.10 M | 146,180 (1,858) | 7.90 M |
|  | M | 10,322 (2,869) | 0.40 M | 8,852 (2,214) | 0.40 M | 514 (2,017) | 0.03 M | 2,256 (928) | 0.20 M | 68,808 (1,708) | 4.00 M | 90,752 (1,795) | 5.10 M |
| All |  | 715,487 (1,655) | 43.30 M | 1,171,781 (1,935) | 60.60 M | 58,998 (2,133) | 2.80 M | 154,517 (744) | 20.80 M | 2,546,159 (1,267) | 200.90 M | 4,646,942 (1,416) | 328.20 M |

Table 2: Logistic regression on CDC COVID case data showing the risk of COVID-19 cases for each age/race/sex group sorted in order by case risk/100,000 U.S. inhabitants

|  | A | B | C | D | E | F | G | H |
| --- | --- | --- | --- | --- | --- | --- | --- | --- |
|  | Race | Age | Sex | No. of Cases | No. in Population^[6]^ | Case risk/100,000 (95% CI^[7]^) | Cum. No. of cases | Cum. No. in population |
| 1 | Am. Indian^[8]^ | 30 - 39 | Female | 5,933 | 0.19 M^[9]^ | 3,163.4 (3,101.0 - 3,226.9) | 5,933 | 0.19 M |
| 2 | Am. Indian | 40 - 49 | Female | 4,735 | 0.16 M | 2,997.2 (2,932.5 - 3,063.2) | 10,668 | 0.35 M |
| 3 | Am. Indian | 20 - 29 | Female | 6,085 | 0.22 M | 2,821.4 (2,766.0 - 2,877.8) | 16,753 | 0.57 M |
| 4 | Hispanic | 40 - 49 | Female | 107,877 | 3.92 M | 2,764.2 (2,750.5 - 2,778.0) | 124,630 | 4.49 M |
| 5 | Hispanic | 20 - 29 | Female | 127,038 | 4.73 M | 2,689.4 (2,677.0 - 2,701.9) | 251,668 | 9.22 M |
| 6 | Black | 80+ | Female | 17,529 | 0.70 M | 2,667.5 (2,635.8 - 2,699.7) | 269,197 | 9.93 M |
| 7 | Hispanic | 30 - 39 | Female | 114,899 | 4.34 M | 2,642.4 (2,629.7 - 2,655.3) | 384,096 | 14.27 M |
| 8 | Am. Indian | 30 - 39 | Male | 4,940 | 0.19 M | 2,624.0 (2,570.4 - 2,678.8) | 389,036 | 14.45 M |
| 9 | Hispanic | 40 - 49 | Male | 104,477 | 4.04 M | 2,572.6 (2,559.6 - 2,585.6) | 493,513 | 18.49 M |
| 10 | Am. Indian | 40 - 49 | Male | 4,126 | 0.16 M | 2,571.0 (2,513.8 - 2,629.4) | 497,639 | 18.65 M |
| 11 | Black | 80+ | Male | 10,322 | 0.36 M | 2,531.6 (2,498.5 - 2,565.2) | 507,961 | 19.01 M |
| 12 | Hispanic | 50 - 59 | Male | 77,349 | 3.08 M | 2,530.9 (2,516.6 - 2,545.3) | 585,310 | 22.09 M |
| 13 | Hispanic | 50 - 59 | Female | 77,743 | 3.08 M | 2,503.7 (2,489.5 - 2,518.0) | 663,053 | 25.17 M |
| 14 | Am. Indian | 50 - 59 | Female | 4,326 | 0.18 M | 2,466.5 (2,410.6 - 2,523.8) | 667,379 | 25.34 M |
| 15 | Hispanic | 30 - 39 | Male | 112,841 | 4.76 M | 2,377.1 (2,365.5 - 2,388.7) | 780,220 | 30.10 M |
| 16 | Am. Indian | 50 - 59 | Male | 3,671 | 0.16 M | 2,297.2 (2,243.9 - 2,351.7) | 783,891 | 30.26 M |
| 17 | Black | 30 - 39 | Female | 72,451 | 3.10 M | 2,286.5 (2,272.6 - 2,300.4) | 856,342 | 33.36 M |
| 18 | Hispanic | 20 - 29 | Male | 114,993 | 5.05 M | 2,271.8 (2,261.0 - 2,282.7) | 971,335 | 38.41 M |
| 19 | Black | 40 - 49 | Female | 62,765 | 2.78 M | 2,248.7 (2,234.3 - 2,263.3) | 1,034,100 | 41.19 M |
| 20 | Hispanic | 60 - 69 | Male | 40,695 | 1.83 M | 2,229.6 (2,212.8 - 2,246.4) | 1,074,795 | 43.02 M |
| 21 | Am. Indian | 20 - 29 | Male | 4,894 | 0.22 M | 2,196.0 (2,151.1 - 2,241.8) | 1,079,689 | 43.24 M |
| 22 | Am. Indian | 80+ | Female | 841 | 0.04 M | 2,159.3 (2,047.6 - 2,277.0) | 1,080,530 | 43.28 M |
| 23 | Black | 50 - 59 | Female | 60,604 | 2.81 M | 2,155.8 (2,141.8 - 2,169.8) | 1,141,134 | 46.09 M |
| 24 | Black | 70 - 79 | Male | 20,068 | 0.89 M | 2,148.3 (2,126.0 - 2,170.8) | 1,161,202 | 46.98 M |
| 25 | White | 20 - 29 | Female | 254,471 | 12.12 M | 2,112.7 (2,105.4 - 2,120.1) | 1,415,673 | 59.10 M |
| 26 | Black | 20 - 29 | Female | 74,736 | 3.43 M | 2,108.4 (2,095.8 - 2,121.1) | 1,490,409 | 62.53 M |
| 27 | Hispanic | 80+ | Male | 8,852 | 0.40 M | 2,090.2 (2,060.3 - 2,120.5) | 1,499,261 | 62.93 M |
| 28 | Am. Indian | 80+ | Male | 514 | 0.03 M | 2,047.5 (1,940.0 - 2,160.7) | 1,499,775 | 62.96 M |
| 29 | Am. Indian | 60 - 69 | Male | 2,503 | 0.13 M | 2,031.8 (1,976.6 - 2,088.6) | 1,502,278 | 63.09 M |
| 30 | Hispanic | 80+ | Female | 12,433 | 0.64 M | 2,030.4 (2,002.7 - 2,058.6) | 1,514,711 | 63.72 M |
| 31 | Hispanic | 70 - 79 | Male | 17,741 | 0.86 M | 2,019.1 (1,996.9 - 2,041.5) | 1,532,452 | 64.59 M |
| 32 | Black | 50 - 59 | Male | 48,846 | 2.44 M | 2,008.5 (1,994.9 - 2,022.3) | 1,581,298 | 67.02 M |
| 33 | Black | 60 - 69 | Male | 38,149 | 1.88 M | 2,003.9 (1,988.4 - 2,019.4) | 1,619,447 | 68.90 M |
| 34 | Am. Indian | 70 - 79 | Male | 1,171 | 0.07 M | 1,954.3 (1,880.5 - 2,030.9) | 1,620,618 | 68.97 M |
| 35 | Hispanic | 60 - 69 | Female | 39,904 | 2.04 M | 1,951.9 (1,937.1 - 1,966.8) | 1,660,522 | 71.00 M |
| 36 | Am. Indian | 60 - 69 | Female | 3,021 | 0.15 M | 1,931.1 (1,879.0 - 1,984.6) | 1,663,543 | 71.16 M |
| 37 | Black | 40 - 49 | Male | 47,022 | 2.46 M | 1,928.1 (1,914.8 - 1,941.4) | 1,710,565 | 73.61 M |
| 38 | Black | 70 - 79 | Female | 23,241 | 1.26 M | 1,919.2 (1,900.0 - 1,938.5) | 1,733,806 | 74.88 M |
| 39 | Black | 60 - 69 | Female | 44,100 | 2.34 M | 1,903.3 (1,889.1 - 1,917.6) | 1,777,906 | 77.22 M |
| 40 | Am. Indian | 10 - 19 | Female | 3,735 | 0.20 M | 1,897.2 (1,851.8 - 1,943.8) | 1,781,641 | 77.42 M |
| 41 | Black | 30 - 39 | Male | 53,717 | 2.91 M | 1,894.9 (1,882.7 - 1,907.2) | 1,835,358 | 80.34 M |
| 42 | White | 80+ | Female | 112,207 | 6.13 M | 1,804.4 (1,794.4 - 1,814.5) | 1,947,565 | 86.47 M |
| 43 | White | 80+ | Male | 68,808 | 4.03 M | 1,748.6 (1,736.7 - 1,760.5) | 2,016,373 | 90.49 M |
| 44 | Am. Indian | 70 - 79 | Female | 1,520 | 0.08 M | 1,746.6 (1,680.7 - 1,815.0) | 2,017,893 | 90.57 M |
| 45 | White | 20 - 29 | Male | 215,143 | 12.72 M | 1,678.2 (1,671.9 - 1,684.5) | 2,233,036 | 103.29 M |
| 46 | Hispanic | 70 - 79 | Female | 17,947 | 1.10 M | 1,661.7 (1,643.5 - 1,680.1) | 2,250,983 | 104.39 M |
| 47 | Black | 20 - 29 | Male | 55,129 | 3.51 M | 1,639.4 (1,629.1 - 1,649.9) | 2,306,112 | 107.90 M |
| 48 | Am. Indian | 10 - 19 | Male | 3,406 | 0.21 M | 1,582.0 (1,543.3 - 1,621.7) | 2,309,518 | 108.11 M |
| 49 | White | 40 - 49 | Female | 176,915 | 11.83 M | 1,493.0 (1,486.9 - 1,499.2) | 2,486,433 | 119.94 M |
| 50 | White | 30 - 39 | Female | 183,972 | 12.46 M | 1,487.3 (1,481.3 - 1,493.3) | 2,670,405 | 132.39 M |
| 51 | White | 50 - 59 | Female | 195,134 | 14.22 M | 1,377.9 (1,372.4 - 1,383.4) | 2,865,539 | 146.62 M |
| 52 | Hispanic | 10 - 19 | Female | 68,694 | 5.08 M | 1,350.0 (1,341.7 - 1,358.4) | 2,934,233 | 151.70 M |
| 53 | White | 50 - 59 | Male | 183,066 | 13.90 M | 1,310.9 (1,305.5 - 1,316.3) | 3,117,299 | 165.60 M |
| 54 | White | 40 - 49 | Male | 155,308 | 11.91 M | 1,306.4 (1,300.7 - 1,312.0) | 3,272,607 | 177.50 M |
| 55 | White | 30 - 39 | Male | 160,821 | 12.69 M | 1,257.5 (1,252.2 - 1,262.9) | 3,433,428 | 190.19 M |
| 56 | White | 10 - 19 | Female | 137,951 | 10.87 M | 1,257.1 (1,251.1 - 1,263.0) | 3,571,379 | 201.06 M |
| 57 | Hispanic | 10 - 19 | Male | 64,451 | 5.28 M | 1,221.9 (1,214.3 - 1,229.6) | 3,635,830 | 206.35 M |
| 58 | White | 70 - 79 | Male | 100,689 | 8.50 M | 1,197.2 (1,190.5 - 1,204.0) | 3,736,519 | 214.85 M |
| 59 | White | 60 - 69 | Male | 153,468 | 13.34 M | 1,151.1 (1,145.9 - 1,156.3) | 3,889,987 | 228.18 M |
| 60 | White | 10 - 19 | Male | 120,966 | 11.42 M | 1,070.4 (1,065.1 - 1,075.7) | 4,010,953 | 239.61 M |
| 61 | White | 60 - 69 | Female | 153,478 | 14.34 M | 1,069.7 (1,064.8 - 1,074.5) | 4,164,431 | 253.95 M |
| 62 | Asian | 20 - 29 | Female | 16,083 | 1.57 M | 1,051.9 (1,039.2 - 1,064.7) | 4,180,514 | 255.52 M |
| 63 | White | 70 - 79 | Female | 103,470 | 9.79 M | 1,045.8 (1,039.9 - 1,051.7) | 4,283,984 | 265.30 M |
| 64 | Black | 10 - 19 | Female | 30,480 | 3.10 M | 1,024.2 (1,015.3 - 1,033.1) | 4,314,464 | 268.41 M |
| 65 | Asian | 20 - 29 | Male | 15,467 | 1.57 M | 959.8 (948.1 - 971.7) | 4,329,931 | 269.98 M |
| 66 | Asian | 80+ | Male | 2,256 | 0.24 M | 959.4 (933.4 - 986.2) | 4,332,187 | 270.22 M |
| 67 | Am. Indian | 0 - 9 | Female | 1,827 | 0.20 M | 913.1 (882.6 - 944.6) | 4,334,014 | 270.42 M |
| 68 | Asian | 50 - 59 | Male | 10,315 | 1.16 M | 912.1 (899.0 - 925.3) | 4,344,329 | 271.57 M |
| 69 | Am. Indian | 0 - 9 | Male | 1,750 | 0.20 M | 884.3 (854.8 - 914.8) | 4,346,079 | 271.77 M |
| 70 | Asian | 80+ | Female | 3,170 | 0.36 M | 860.2 (837.3 - 883.8) | 4,349,249 | 272.13 M |
| 71 | Asian | 30 - 39 | Female | 15,356 | 1.84 M | 853.9 (843.4 - 864.5) | 4,364,605 | 273.97 M |
| 72 | Black | 10 - 19 | Male | 28,505 | 3.19 M | 853.3 (845.7 - 861.0) | 4,393,110 | 277.17 M |
| 73 | Asian | 60 - 69 | Male | 7,267 | 0.88 M | 849.1 (834.9 - 863.6) | 4,400,377 | 278.05 M |
| 74 | Asian | 50 - 59 | Female | 11,349 | 1.34 M | 832.9 (821.1 - 844.9) | 4,411,726 | 279.38 M |
| 75 | Asian | 30 - 39 | Male | 14,458 | 1.70 M | 830.5 (820.1 - 841.1) | 4,426,184 | 281.08 M |
| 76 | Asian | 40 - 49 | Male | 11,394 | 1.43 M | 801.4 (790.5 - 812.5) | 4,437,578 | 282.52 M |
| 77 | Asian | 40 - 49 | Female | 13,098 | 1.63 M | 796.1 (785.5 - 806.9) | 4,450,676 | 284.15 M |
| 78 | Asian | 70 - 79 | Male | 3,572 | 0.50 M | 735.1 (717.8 - 752.8) | 4,454,248 | 284.65 M |
| 79 | Asian | 60 - 69 | Female | 7,713 | 1.09 M | 685.1 (673.6 - 696.8) | 4,461,961 | 285.74 M |
| 80 | Hispanic | 0 - 9 | Male | 32,740 | 5.27 M | 633.1 (627.4 - 638.9) | 4,494,701 | 291.01 M |
| 81 | Hispanic | 0 - 9 | Female | 31,107 | 5.07 M | 601.4 (595.9 - 607.1) | 4,525,808 | 296.08 M |
| 82 | Asian | 70 - 79 | Female | 3,619 | 0.63 M | 557.1 (544.0 - 570.6) | 4,529,427 | 296.71 M |
| 83 | Asian | 10 - 19 | Female | 6,504 | 1.24 M | 529.3 (519.8 - 538.9) | 4,535,931 | 297.95 M |
| 84 | Asian | 10 - 19 | Male | 6,559 | 1.26 M | 518.6 (509.3 - 528.1) | 4,542,490 | 299.20 M |
| 85 | Black | 0 - 9 | Female | 13,654 | 2.99 M | 465.2 (459.3 - 471.2) | 4,556,144 | 302.19 M |
| 86 | Black | 0 - 9 | Male | 14,169 | 3.09 M | 450.8 (445.0 - 456.6) | 4,570,313 | 305.28 M |
| 87 | White | 0 - 9 | Female | 33,967 | 10.06 M | 342.4 (339.4 - 345.4) | 4,604,280 | 315.34 M |
| 88 | White | 0 - 9 | Male | 36,325 | 10.58 M | 339.0 (336.0 - 342.0) | 4,640,605 | 325.91 M |
| 89 | Asian | 0 - 9 | Male | 3,410 | 1.19 M | 290.0 (282.7 - 297.4) | 4,644,015 | 327.11 M |
| 90 | Asian | 0 - 9 | Female | 2,927 | 1.13 M | 254.2 (247.7 - 260.8) | 4,646,942 | 328.24 M |

Table 3: Predictive Coefficients (Parameter Estimates) Of Maximum Likelihood of Death from SAS Version 9.4 PROC Logistic

| **Parameter** | | | **Coefficient/**  **Estimate** | **Standard Error** | **95% Confidence**  **Limits (Upper)** | **95% Confidence**  **Limits (Lower)** | **Pr > ChiSq** |
| --- | --- | --- | --- | --- | --- | --- | --- |
| Intercept |  |  | -22.1135 | 21.9719 | -65.1777 | 20.9507 | 0.3142 |
| AGE | 15-24 years |  | 8.8154 | 21.9726 | -34.2502 | 51.881 | 0.6883 |
| AGE | 25-34 years |  | 10.9359 | 21.972 | -32.1285 | 54.0002 | 0.6187 |
| AGE | 35-44 years |  | 11.8438 | 21.972 | -31.2204 | 54.9081 | 0.5899 |
| AGE | 45-54 years |  | 13.1234 | 21.9719 | -29.9409 | 56.1876 | 0.5503 |
| AGE | 5-14 years |  | -0.1249 | 27.2587 | -53.551 | 53.3012 | 0.9963 |
| AGE | 55-64 years |  | 14.2488 | 21.9719 | -28.8155 | 57.313 | 0.5167 |
| AGE | 65-74 years |  | 15.3395 | 21.9719 | -27.7247 | 58.4037 | 0.4851 |
| AGE | 75-84 years |  | 16.4747 | 21.9719 | -26.5895 | 59.5389 | 0.4534 |
| AGE | 85 years and over |  | 17.7304 | 21.9719 | -25.3338 | 60.7946 | 0.4197 |
| RACE | American Indian |  | 0.0323 | 158.2 | -310 | 310.1 | 0.9998 |
| RACE | Asian |  | -0.0432 | 70.793 | -138.8 | 138.7 | 0.9995 |
| RACE | Black |  | 0.0631 | 44.9886 | -88.1129 | 88.2392 | 0.9989 |
| RACE | Hispanic |  | 0.0225 | 37.7575 | -73.9809 | 74.0258 | 0.9995 |
| SEX | Female |  | -0.4659 | 0.00433 | -0.4744 | -0.4574 | <.0001 |
| RACE*AGE | American Indian | 15-24 years | -9.0567 | 199.1 | -399.3 | 381.2 | 0.9637 |
| RACE*AGE | American Indian | 25-34 years | 1.9996 | 158.2 | -308.1 | 312.1 | 0.9899 |
| RACE*AGE | American Indian | 35-44 years | 2.3121 | 158.2 | -307.7 | 312.4 | 0.9883 |
| RACE*AGE | American Indian | 45-54 years | 2.1217 | 158.2 | -307.9 | 312.2 | 0.9893 |
| RACE*AGE | American Indian | 5-14 years | -0.117 | 200.2 | -392.5 | 392.2 | 0.9995 |
| RACE*AGE | American Indian | 55-64 years | 1.5385 | 158.2 | -308.5 | 311.6 | 0.9922 |
| RACE*AGE | American Indian | 65-74 years | 1.103 | 158.2 | -309 | 311.2 | 0.9944 |
| RACE*AGE | American Indian | 75-84 years | 0.6249 | 158.2 | -309.4 | 310.7 | 0.9968 |
| RACE*AGE | American Indian | 85 years and over | -0.0526 | 158.2 | -310.1 | 310 | 0.9997 |
| RACE*AGE | Asian | 15-24 years | -9.0438 | 86.2115 | -178 | 159.9 | 0.9165 |
| RACE*AGE | Asian | 25-34 years | -1.009 | 70.7935 | -139.8 | 137.7 | 0.9886 |
| RACE*AGE | Asian | 35-44 years | 0.1806 | 70.793 | -138.6 | 138.9 | 0.998 |
| RACE*AGE | Asian | 45-54 years | 0.4518 | 70.793 | -138.3 | 139.2 | 0.9949 |
| RACE*AGE | Asian | 5-14 years | -0.107 | 89.1143 | -174.8 | 174.6 | 0.999 |
| RACE*AGE | Asian | 55-64 years | 0.5373 | 70.793 | -138.2 | 139.3 | 0.9939 |
| RACE*AGE | Asian | 65-74 years | 0.3755 | 70.793 | -138.4 | 139.1 | 0.9958 |
| RACE*AGE | Asian | 75-84 years | 0.0857 | 70.793 | -138.7 | 138.8 | 0.999 |
| RACE*AGE | Asian | 85 years and over | -0.0773 | 70.793 | -138.8 | 138.7 | 0.9991 |
| RACE*AGE | Black | 15-24 years | 2.0809 | 44.9891 | -86.0961 | 90.258 | 0.9631 |
| RACE*AGE | Black | 25-34 years | 1.7023 | 44.9887 | -86.4738 | 89.8785 | 0.9698 |
| RACE*AGE | Black | 35-44 years | 1.956 | 44.9886 | -86.2201 | 90.1321 | 0.9653 |
| RACE*AGE | Black | 45-54 years | 1.678 | 44.9886 | -86.4981 | 89.8541 | 0.9702 |
| RACE*AGE | Black | 5-14 years | -0.1223 | 56.8567 | -111.6 | 111.3 | 0.9983 |
| RACE*AGE | Black | 55-64 years | 1.5199 | 44.9886 | -86.6561 | 89.696 | 0.973 |
| RACE*AGE | Black | 65-74 years | 1.3472 | 44.9886 | -86.8288 | 89.5233 | 0.9761 |
| RACE*AGE | Black | 75-84 years | 0.9955 | 44.9886 | -87.1806 | 89.1715 | 0.9823 |
| RACE*AGE | Black | 85 years and over | 0.5071 | 44.9886 | -87.669 | 88.6831 | 0.991 |
| RACE*AGE | Hispanic | 15-24 years | 2.2452 | 37.758 | -71.7591 | 76.2496 | 0.9526 |
| RACE*AGE | Hispanic | 25-34 years | 1.8538 | 37.7576 | -72.1496 | 75.8573 | 0.9608 |
| RACE*AGE | Hispanic | 35-44 years | 2.1361 | 37.7575 | -71.8673 | 76.1395 | 0.9549 |
| RACE*AGE | Hispanic | 45-54 years | 1.9088 | 37.7575 | -72.0946 | 75.9121 | 0.9597 |
| RACE*AGE | Hispanic | 5-14 years | -0.1171 | 47.5909 | -93.3935 | 93.1594 | 0.998 |
| RACE*AGE | Hispanic | 55-64 years | 1.7204 | 37.7575 | -72.2829 | 75.7238 | 0.9637 |
| RACE*AGE | Hispanic | 65-74 years | 1.4479 | 37.7575 | -72.5554 | 75.4512 | 0.9694 |
| RACE*AGE | Hispanic | 75-84 years | 0.984 | 37.7575 | -73.0193 | 74.9873 | 0.9792 |
| RACE*AGE | Hispanic | 85 years and over | 1 | 0.4438 | 37.7575 | -73.5596 | 74.4471 |

Table 4: Predictive Coefficients (Parameter Estimates) Of Maximum Likelihood of Case from SAS PROC from SAS Version 9.4 PROC Logistic

| **Parameter** |  |  | **Coefficient/**  **Estimate** | **Standard Error** | **95% Confidence**  **Limits (Upper)** | **95% Confidence**  **Limits (Lower)** | **Pr > ChiSq** |
| --- | --- | --- | --- | --- | --- | --- | --- |
| **Intercept** |  |  | -5.6835 | 0.0045 | -5.6924 | -5.6747 | <.0001 |
| **AGE** | 10 - 19 Years |  | 1.1572 | 0.0051 | 1.1472 | 1.1672 | <.0001 |
| **AGE** | 20 - 29 Years |  | 1.613 | 0.0048 | 1.6035 | 1.6225 | <.0001 |
| **AGE** | 30 - 39 Years |  | 1.3202 | 0.0049 | 1.3105 | 1.3299 | <.0001 |
| **AGE** | 40 - 49 Years |  | 1.3588 | 0.005 | 1.349 | 1.3685 | <.0001 |
| **AGE** | 50 - 59 Years |  | 1.3623 | 0.0049 | 1.3526 | 1.3719 | <.0001 |
| **AGE** | 60 - 69 Years |  | 1.2307 | 0.005 | 1.2208 | 1.2405 | <.0001 |
| **AGE** | 70 - 79 Years |  | 1.2704 | 0.0053 | 1.26 | 1.2809 | <.0001 |
| **AGE** | 80+ Years |  | 1.6548 | 0.0057 | 1.6437 | 1.666 | <.0001 |
| **RACE** | Black |  | 0.2861 | 0.0072 | 0.272 | 0.3003 | <.0001 |
| **RACE** | Hispanic |  | 0.6276 | 0.0056 | 0.6167 | 0.6386 | <.0001 |
| **RACE** | American Indian |  | 0.9643 | 0.0177 | 0.9296 | 0.9991 | <.0001 |
| **RACE** | Asian |  | -0.1567 | 0.0134 | -0.1828 | -0.1305 | <.0001 |
| **sex** | Female |  | 0.01 | 0.005 | 0.0002 | 0.0197 | 0.0445 |
| **RACE*AGE** | Black | 10 - 19 Years | -0.515 | 0.0085 | -0.5316 | -0.4984 | <.0001 |
| **RACE*AGE** | Black | 20 - 29 Years | -0.3099 | 0.0078 | -0.3251 | -0.2947 | <.0001 |
| **RACE*AGE** | Black | 30 - 39 Years | 0.1304 | 0.0078 | 0.115 | 0.1458 | <.0001 |
| **RACE*AGE** | Black | 40 - 49 Years | 0.1095 | 0.0079 | 0.0939 | 0.125 | <.0001 |
| **RACE*AGE** | Black | 50 - 59 Years | 0.1477 | 0.0079 | 0.1322 | 0.1632 | <.0001 |
| **RACE*AGE** | Black | 60 - 69 Years | 0.2769 | 0.0081 | 0.261 | 0.2929 | <.0001 |
| **RACE*AGE** | Black | 70 - 79 Years | 0.3082 | 0.0089 | 0.2908 | 0.3256 | <.0001 |
| **RACE*AGE** | Black | 80+ Years | 0.092 | 0.0097 | 0.073 | 0.1109 | <.0001 |
| **RACE*AGE** | Hispanic | 10 - 19 Years | -0.4937 | 0.0064 | -0.5064 | -0.4811 | <.0001 |
| **RACE*AGE** | Hispanic | 20 - 29 Years | -0.3188 | 0.006 | -0.3306 | -0.3069 | <.0001 |
| **RACE*AGE** | Hispanic | 30 - 39 Years | 0.0205 | 0.0061 | 0.0085 | 0.0325 | 0.0008 |
| **RACE*AGE** | Hispanic | 40 - 49 Years | 0.0629 | 0.0062 | 0.0509 | 0.075 | <.0001 |
| **RACE*AGE** | Hispanic | 50 - 59 Years | 0.0427 | 0.0063 | 0.0304 | 0.055 | <.0001 |
| **RACE*AGE** | Hispanic | 60 - 69 Years | 0.0444 | 0.0068 | 0.0311 | 0.0577 | <.0001 |
| **RACE*AGE** | Hispanic | 70 - 79 Years | -0.0967 | 0.008 | -0.1123 | -0.081 | <.0001 |
| **RACE*AGE** | Hispanic | 80+ Years | -0.4457 | 0.0091 | -0.4636 | -0.4278 | <.0001 |
| **RACE*AGE** | American Indian | 10 - 19 Years | -0.5685 | 0.021 | -0.6098 | -0.5273 | <.0001 |
| **RACE*AGE** | American Indian | 20 - 29 Years | -0.6901 | 0.0198 | -0.7289 | -0.6513 | <.0001 |
| **RACE*AGE** | American Indian | 30 - 39 Years | -0.2148 | 0.0199 | -0.2537 | -0.1759 | <.0001 |
| **RACE*AGE** | American Indian | 40 - 49 Years | -0.2744 | 0.0204 | -0.3144 | -0.2344 | <.0001 |
| **RACE*AGE** | American Indian | 50 - 59 Years | -0.3933 | 0.0207 | -0.4338 | -0.3528 | <.0001 |
| **RACE*AGE** | American Indian | 60 - 69 Years | -0.3871 | 0.022 | -0.4303 | -0.344 | <.0001 |
| **RACE*AGE** | American Indian | 70 - 79 Years | -0.4666 | 0.0261 | -0.5177 | -0.4155 | <.0001 |
| **RACE*AGE** | American Indian | 80+ Years | -0.8035 | 0.0325 | -0.8672 | -0.7398 | <.0001 |
| **RACE*AGE** | Asian | 10 - 19 Years | -0.5736 | 0.0159 | -0.6048 | -0.5423 | <.0001 |
| **RACE*AGE** | Asian | 20 - 29 Years | -0.4094 | 0.0144 | -0.4376 | -0.3812 | <.0001 |
| **RACE*AGE** | Asian | 30 - 39 Years | -0.2625 | 0.0145 | -0.2909 | -0.2342 | <.0001 |
| **RACE*AGE** | Asian | 40 - 49 Years | -0.337 | 0.0147 | -0.3659 | -0.3082 | <.0001 |
| **RACE*AGE** | Asian | 50 - 59 Years | -0.2101 | 0.0149 | -0.2393 | -0.1809 | <.0001 |
| **RACE*AGE** | Asian | 60 - 69 Years | -0.1507 | 0.0156 | -0.1812 | -0.1201 | <.0001 |
| **RACE*AGE** | Asian | 70 - 79 Years | -0.3358 | 0.0178 | -0.3707 | -0.3009 | <.0001 |
| **RACE*AGE** | Asian | 80+ Years | -0.4516 | 0.0191 | -0.489 | -0.4141 | <.0001 |
| **AGE*sex** | 10 - 19 Years | Female | 0.1526 | 0.0057 | 0.1415 | 0.1637 | <.0001 |
| **AGE*sex** | 20 - 29 Years | Female | 0.2247 | 0.0053 | 0.2143 | 0.2351 | <.0001 |
| **AGE*sex** | 30 - 39 Years | Female | 0.1602 | 0.0054 | 0.1496 | 0.1707 | <.0001 |
| **AGE*sex** | 40 - 49 Years | Female | 0.1255 | 0.0054 | 0.1148 | 0.1361 | <.0001 |
| **AGE*sex** | 50 - 59 Years | Female | 0.0406 | 0.0054 | 0.0299 | 0.0512 | <.0001 |
| **AGE*sex** | 60 - 69 Years | Female | -0.0842 | 0.0057 | -0.0953 | -0.0731 | <.0001 |
| **AGE*sex** | 70 - 79 Years | Female | -0.1468 | 0.0061 | -0.1588 | -0.1347 | <.0001 |
| **AGE*sex** | 80+ Years | Female | 0.022 | 0.0065 | 0.0093 | 0.0348 | 0.0007 |
| **RACE*sex** | Black | Female | 0.0217 | 0.0027 | 0.0163 | 0.027 | <.0001 |
| **RACE*sex** | Hispanic | Female | -0.0616 | 0.0023 | -0.0661 | -0.0571 | <.0001 |
| **RACE*sex** | American Indian | Female | 0.0223 | 0.0085 | 0.0057 | 0.0389 | 0.0084 |
| **RACE*sex** | Asian | Female | -0.1422 | 0.0053 | -0.1525 | -0.1318 | <.0001 |

1. American Indian
2. No. in population calculated based on NCHS bridged race-ethnicity population estimates
3. Millions
4. Female
5. Male
6. No. in population calculated based on NCHS bridged race-ethnicity population estimates
7. Confidence Interval
8. American Indian
9. Millions
